# Is a 14-day quarantine period optimal for effectively controlling coronavirus disease 2019 (COVID-19)?

**DOI:** 10.1101/2020.03.15.20036533

**Authors:** Xue Jiang, Yawei Niu, Xiong Li, Lin Li, Wenxiang Cai, Yucan Chen, Bo Liao, Edwin Wang

## Abstract

**Background:** The outbreak of a new coronavirus (SARS-CoV-2) disease (Covid-19) has become pandemic. To be more effectively controlling the disease, it is critical to set up an optimal quarantine period so that about 95% of the cases developing symptoms will be retained for isolation. At the moment, the WHO-established quarantine period is 14 days based on previous reports which had studied small sizes of hospitalized cases (10 and ∼100, respectively), however, over 80% of adult- and 95% of child-cases were not necessary to stay in hospitals, and therefore, had not been hospitalized. Therefore, we are questioning if the current-inferred median incubation time is representative for the whole Covid-19 population, and if the current quarantine period is optimal.

**Methods:** We compiled and analyzed the patient-level information of 2015 laboratory-confirmed Covid-19 cases including 99 children in 28 Chinese provinces. This cohort represents a wide-range spectrum of Covid-19 disease with both hospitalized and non-hospitalized cases.

**Results:** The full range of incubation periods of the Covid-19 cases ranged from 0 to 33 days among 2015 cases. There were 6 (0.13%) symptom-free cases including 4 females with a median age of 25.5 years and 2 males with a median age of 36 years. The median incubation period of both male and female adults was similar (7-day) but significantly shorter than that (9-day) of child cases (P=0.02). This cohort contained 4 transmission generations, and incubation periods of the cases between generations were not significantly different, suggesting that the virus has not been rapidly adapted to human beings. Interestingly, incubation periods of 233 cases (11.6%) were longer than the WHO-established quarantine period (14 days). Data modeling suggested that if adults take an extra 4-day or 7-day of isolation (i.e., a quarantine period of 18 or 21 days), 96.2% or 98.3%, respectively, of the people who are developing symptoms will be more effectively quarantined. Patients transmitted via lunch/dinner parties (i.e., gastrointestinal tract infection through oral transmission) had a significantly longer incubation period (9-day) than other adults transmitted via respiratory droplets or contaminated surfaces and objects (P<0.004).

**Conclusions:** The whole Covid-19 population including both hospitalized and non-hospitalized cases had a median incubation period of 7-day for adults, which is 1.8-day longer than the hospitalized cases reported previously. An extension of the adult quarantine period to 18 days or 21 days could be more effective in preventing virus-spreading and controlling the disease. The cases transmitted by lunch/dinner parties could be infected first in the gastrointestinal tract through oral transmission and then infected in the respiratory system so that they had a longer incubation period.

## Introduction

The outbreak of a new coronavirus (SARS-CoV-2) disease (Covid-19) has become a worldwide pandemic. Understanding the characteristics of the incubation period is crucial to design public health efforts for effectively isolating the cases and contacts to control outbreaks of Covid-19. SARS-CoV-2 can be transmitted by either respiratory droplets, contaminated surfaces, and objects or an oral-fecal transmission route to lead gastrointestinal tract infection^1^. Further, the virus can survive on surfaces for up to several days^2^. These facts suggested that SARS-CoV-2 may have different clinical characteristics than other coronaviruses.

Because outbreaks are being occurred in many countries, it is critical to analyze real-time data of the Covid-19 cases to gain insights to inform public-health intervention strategies for managing and preventing the disease. One of the key issues for controlling Covid-19 is to set up a suitable quarantine period which could capture about 95% of the cases developing symptoms so that the practice of isolations will be more effective and save public health resources. Thus far, the incubation time studies have been focused on hospitalized patients who were highly selected based on medical conditions, and further, sample sizes for inferring incubation times were small, for example, 10 and ∼100, respectively^3,4^. However, the majority (i.e., about 80% of adults and 95% of children) of the Covid-19 cases were not necessary to be hospitalized, and therefore, had not been hospitalized due to their mild conditions. Here, we asked if the median incubation time inferred from the hospitalized patients are representative for the whole population of Covid-19, and if the current quarantine period determined based on hospitalized patients is optimal.

To answer these questions, we manually curated the patient-level data from the case reports on official websites of China’s provincial public health agencies. This cohort represents a general population of Covid-19 cases without a selection based on medical conditions. The cohort contained 2015 cases including 99 children who had symptom onsets between January 1, 2020, and February 25, 2020. All the curated cases contained exposure, onset, and diagnosis dates. We analyzed the incubation time for the Covid-19 population containing both hospitalized and non-hospitalized cases.

## Methods

### Data curation

We manually curated clinical data of the Covid-19 patients from official websites of local Chinese health agencies (Supplementary Table 1) where the Covid-19 case reports have been carefully collected and generated by public health professionals and social workers. All the Covid-19 cases have been laboratory-test confirmed. To extract the individual-level patient data, we read the case reports in Chinese to curate age, sex, demographic information (i.e., name of the provinces and cities), dates of exposures, symptom onset and diagnosis (i.e., confirmed the SARS-CoV-2 infection through laboratory-tests), when available. The patient data were curated by XJ, WXC, and YCC who are native Chinese speakers and translated the summary of each patient into English. To make sure the data accuracy, the curations have been cleaned and checked 4 times by four other researchers, YWN, XL, LL, and EW. We mainly focused on the cases which were allowed to inferring incubation time. Among 4567 cases, after removing the cases which did not have exposure dates, or whose exposure dates were not sure, we obtained 2015 cases whose incubation times could be inferred. Because the information of Covid-19 cases in Hubei province including Wuhan, the city at the center of the epidemic, was not publicly available, this cohort included only non-Hubei cases.

## Study design and data

We curated the patient-level data as described above and characterized patient baseline information such as incubation periods. All the data in this study have been de-identified and publicly available through local Chinese health agencies, therefore, patient consent and ethics approval was not required.

### Statistical analysis

The incubation period of a Covid-19 case is the time between infectious exposure and symptom onset. Over 2,000 cases had sufficient information for inferring incubation periods. Most of which had clear records for the dates of exposures and symptom onsets. For imported cases (e.g., travelers from Wuhan city), the exposure dates were defined as the dates of leaving the epicentral cities such as Wuhan. For the cases, who had been quarantined and monitored for the disease, the diagnosed dates were set as the dates for their symptom onsets. χ2 tests and t-tests were used for statistical analysis using R, a free software environment for statistical computing. We considered p values of less than 0.05 to be significant.

## RESULTS

We collected patient-level data for 2015 Covid-19 cases reported between January 1 and February 25, 2020 (Table 1, Supplementary Table 1). 887 (44.0%) cases were female adults with a median age of 46 years (IQR 41.5-50.5), 1029 (51.0%) were male adults and the median age was 45 years (IQR 40.5-50.5), and 99 (4.9 %) cases were children with the median age was 9 years (IQR 7-11) with a boy-to-girl ratio of 1.3:1. 774 (38.4%) were imported cases, most reported a travel history to Wuhan (133, 17.2%) or were residents of Wuhan (641, 82.8%), while 240 (11.9%) of the imported case were either the residents or returning travelers from non-Wuhan regions.

**Table 1.** Characteristics and social contacts of Covid-19 cases in this study

|  |  |  |  |  |  |  |
| --- | --- | --- | --- | --- | --- | --- |
| Social contact | **Male (N = 1029)** |  | **Female (N = 887)** |  | **Child (N = 99)** |  |
|  | Median age (IQR)<br>yr | Distribution<br>no./total no. (%) | Median age (IQR)<br>yr | Distribution<br>no./total no. (%) | Median age (IQR)<br>yr | Distribution<br>no./total no.<br>(%) |
| Wuhan resident | 41.0 (37.0–45.0) | 381/1029 (0.37) | 45.0 (45.0–4.75) | 293/887 (0.33) | 12.0 (10.0–14.0) | 27/99 (0.27) |
| Traveler back from<br>Wuhan | 40.5 (37.5–43.5) | 82/1029 (0.08) | 43.0 (43.0–5.01) | 53/887 (0.06) | 11.5 (9.5–13.5) | 6/99 (0.06) |
| non-Wuhan region<br>resident | 43.0 (38.5–47.5) | 72/1029 (0.07) | 43.0 (43.0–5.25) | 44/887 (0.05) | 10.0 (7.0–13.0) | 7/99 (0.07) |
| Traveler back from<br>non-Wuhan regions | 45.0 (41.7–48.3) | 82/1029 (0.08) | 41.0 (41.0–3.0) | 53/887 (0.06) | 6.0 (4.0–8.0) | 7/99 (0.07) |
| Local-acquired<br>cases | 50.0 (45.5–55.5) | 412/2095 (0.40) | 52.0 (52.0–3.75) | 443/887 (0.50) | 8.0 (6.2–9.75) | 52/99 (0.53) |

The incubation period is essential in the control of infectious diseases. To halt a pandemic, a suitable quarantine period should be set by investigating incubation periods. Thus, understanding the characteristics of the incubation period is crucial to design public health efforts. In this cohort, the full range of incubation periods of the Covid-19 cases ranged from 0 to 33 days among 2015 cases. Both male and female adults had their median incubation periods of 7-day, which is 1.8-day longer than those in other reports^3,4^ (see discussion). However, a similar result was released in the recent news from the Chinese Medical Association that the median incubation periods were 5-7 days^5^. The median incubation period (9-day) of children were significantly longer than those of adults (t-test P = 0.02). For adults, the incubation periods for the majority (66.5%) of the cases were 1-9 days: most of them (57.6% and 52.0% for males and females, respectively) had 1-5 days incubation.

Importantly, we found that the incubation periods of 11.5% (n=233, 123 and 105 were male and female, respectively) of the cases were longer than 14 days, the current WHO-set 14-day quarantine period. These results agree with a recent study which also reported 12.7% (n=13) of the cases whose incubation periods were longer than 14 days^4^. These data suggested that the official 14-day quarantine period only captured 88.5% of the population developing Covid-19. However, among the 233 cases with longer incubation periods, most (n=198, 85.0%) had 15-21 days of incubation periods, suggesting that implementation of the 21-day quarantine period for everybody will capture 98.3% of the cases. However, we knew that fewer children were infected, we plotted the age-distribution of the cases having a longer quarantine period (Fig 1). Based on Fig 1, we concluded that if adults take an extra 7 days for quarantine, we will capture 98.3% of the cases. Data modeling also showed that a 17-day or 18-day quarantine period will capture 94.8% and 96.2%, respectively, of the cases.

**Fig 1.**
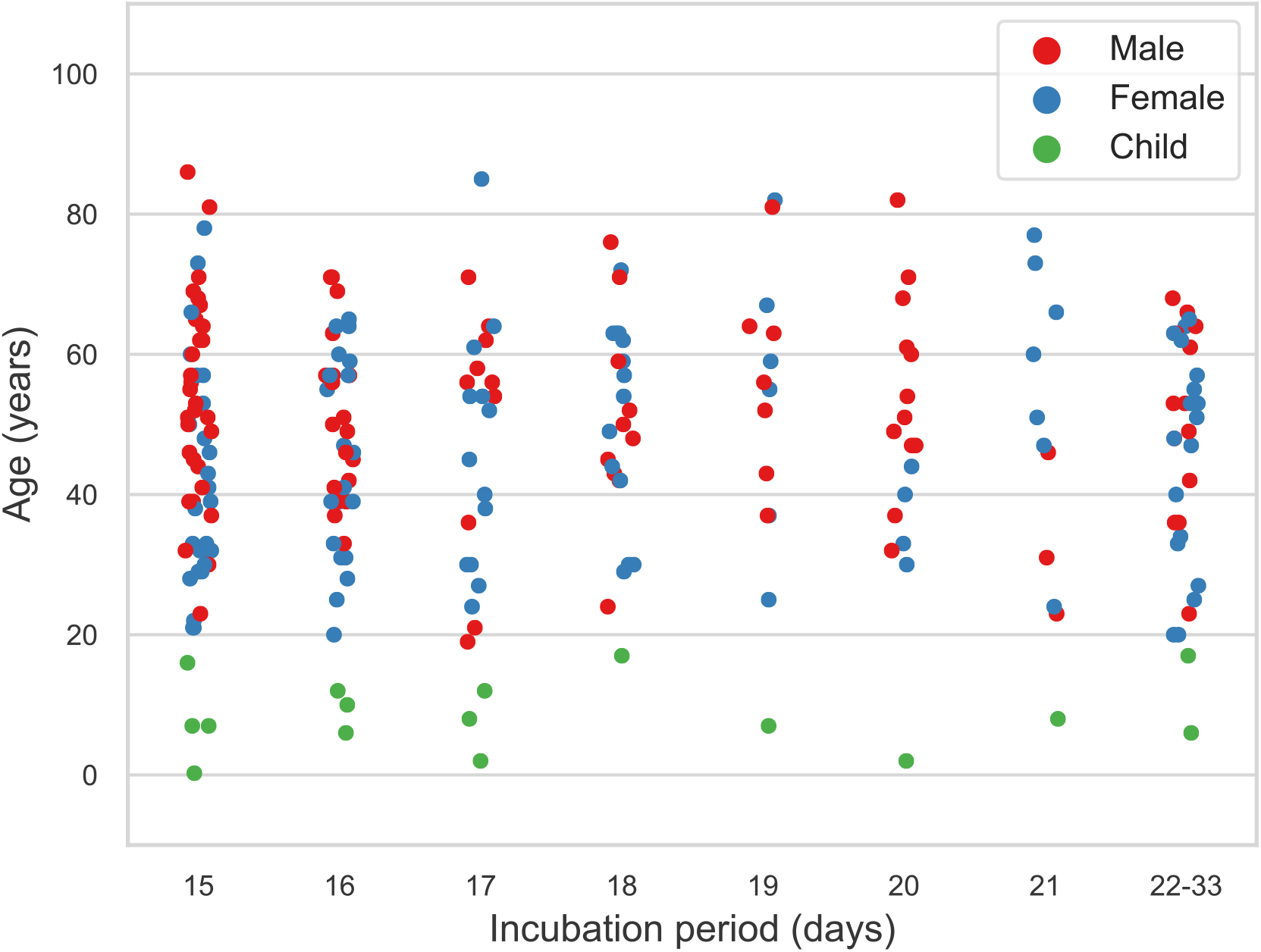
The distribution of the cases having a longer incubation period. The figure shows the ages of the cases who had the incubation days (15-day, 16-day and so on) which were longer than 14 days. Red, blue and green represent men, women, and children, respectively.

The incubation period of 2.0% (n=19 and 20 for male and female adults, respectively) of the total cases was 0-day, while some cases were transmitted by having contacts with Covid-19 patients who had not developed symptom onset yet, suggesting that SARS-CoV-2 might be contagious (i.e., is shedding virus) during incubation periods, and these patients could be more susceptible to this virus. There were 6 (0.13%) symptom-free cases including 4 females with a median age of 25.5 years (IQR 24.0-27.0) and 2 males with a median age of 36 years (IQR 24.0-48.0). These symptom-free cases were much younger than the majority of the patients.

The fecal-oral transmission has been found in Covide-19 cases^1^. Food sharing in lunch/dinner parties could facilitate fecal-oral transmission. Interestingly, the incubation time (9-day IQR 7.75-10.25) of the cases transmitted by lunch/dinner party contacts was significantly longer than others (P=0.03), suggesting that oral transmission via sharing food had longer incubation periods than transmitting via respiratory droplets.

There were 4 generations of transmitted cases based on contact tracing information. The Wuhan-imported cases were defined as the 1-generation cases, while the 2-generation cases included both non-Wuhan imported cases, and new cases which were transmitted directly by a 1-generation case or an unknown cause. The 3-generation cases were the ones transmitted directly by a 2-generation case. The incubation time was not significantly different between the patients of transmission generations, suggesting that the virus is not rapidly adapted to humans.

## Discussion

The incubation period is essential in making intervention strategies to control infectious diseases. Thus, understanding of incubation time of a wide-range spectrum of Covid-19 for establishing an optimal quarantine period is crucial to designing public health interventions. Two recent studies have reported incubation periods for Covid-19 cases^3,4^, but only for the hospitalized patients, who were highly selected and represented only 20% of adult- and 5% of child-cases of the general Covid-19 population. Further, the sample size (about 100 and 10 cases, respectively) for inferring incubation periods were small in these studies. Here, we manually curated a Covid-19 cohort containing baseline and incubation information of nearly 2,000 patients including 99 children. This cohort included both hospitalized and non-hospitalized cases. By analyzing this cohort, we characterized the incubation times of Covid-19 adults and children as well, and examined the effectiveness of the 14-day quarantine period.

The median incubation period for adults in this cohort was 7-day, which was longer than 5-5.2 days reported previously^3,4^. Recently, the Chinese Medical Association also found that the coronavirus median incubation period was 5-7 days^5^. Kids had an incubation period of 9-day and significantly longer than adults. Based on previous studies, WHO has set a 14-day quarantine period for Covid-19. However, we found that that the incubation periods of 11.5% (n=233) of the cases were longer than 14 days, meaning that the current quarantine policy only stops 88% of the cases for virus spreading. This finding is similar to another report showing that 13 (12.7%) cases had their incubation time longer than 14 days^4^. In general, a quarantine period that retains about 95% of the cases developing symptoms will be more effective^6^. Thus, we further analyzed the subset (n=233) and found that 98.7% and 97.9%, respectively, of the men and women cases who are developing the disease, will be quarantined, if everybody or adults alone, respectively, take an extra 7-day of isolation (i.e., a quarantine period of 21 days). If taking a quarantine period of 17-day or 18-day, 94.8% and 96.2%, respectively, of the cases developing symptoms will be successfully quarantined.

As the outbreak is fast-moving in the world, based on this analysis we recommended that an extension of adults’ quarantine period to 17 or 21 days could be more effective. Of note, the outbreak occurred during the Chinese New Year holiday, further the holiday period had been extended to another 10 days^7^. Moreover, until now (as of March 15, 2020) schools and universities have been still shut down. Thus, due to the implementation of the aggressive social distancing measures many people in China have been isolated for more than 60 days.

The full range of incubation periods of the Covid-19 cases ranged from zero to 33 days in this study, including ∼30 cases whose incubation periods ranged 22-33 days, and 6 symptom-free cases, suggesting that a wide-range heterogeneous response to the SARS-CoV-2 virus in the population, and the challenges for preventing virus-spreading. Furthermore, some cases start to transmit this virus before the symptom onset. This is very different from the transmission patterns of SARS (Severe Acute Respiratory Syndrome), for which transmission rarely occurred until after the 4-5 days after symptom onset^8^. When a virus has infectiousness before symptom onset, control of outbreaks using contact tracing and isolation is more challenging^9^.

Interestingly, the cases transmitted by lunch/dinner party contacts had a 2-day longer incubation period than the cases transmitted via respiratory droplets. The Chinese style dish- and food-sharing during lunch/dinner parties can easily contaminant the shared dishes by virus-carriers and thus, could facilitate oral transmission for leading gastrointestinal tract infection, which has been reported as a transmission means for Covide-19^1^. It has been reported that 10% of patients presented with diarrhea and nausea 1-2 days before the development of fever and respiratory symptoms^10,11^. These observations explained why the cases transmitted by lunch/dinner party contacts had significantly longer incubation periods. Therefore, the early signs of the cases transmitted by lunch/dinner parties could be diarrhea and nausea which often develop before fever and respiratory symptoms in these patients.

### Limitations

Several limitations of this study are the following: (1) the cohort did not include patient-level information from the patients in Hubei province including Wuhan city, because they were exhaust-dealing with the outbreak and could not make the information in their official websites. (2) all the cases are Chinese patients.

## Data Availability

All the data in this study have been de-identified and publicly available through local Chinese health agencies, therefore, patient consent and ethics approval was not required.

## Acknowledgments

EW and XJ designed the project. XJ, WXC, and YCC manually curated data and translated the summary of each patient into English, while YWN, XL, LL, and EW checked the curations and cleaned the data. XJ and YWN conducted data analysis. EW and XJ wrote the manuscript. This work is supported by the AISH Translational Chair Program in Cancer Genomics.

## References

1. Yeo C, Kaushal S, Yeo D. Enteric involvement of coronaviruses: is faecal-oral transmission of SARS-CoV-2 possible? Lancet Gastroenterol Hepatol 2020.

2. Kampf G, Todt D, Pfaender S, Steinmann E. Persistence of coronaviruses on inanimate surfaces and their inactivation with biocidal agents. J Hosp Infect 2020;104:246–51.

3. Li Q, Guan X, Wu P, et al. Early Transmission Dynamics in Wuhan, China, of Novel Coronavirus-Infected Pneumonia. N Engl J Med 2020.

4. Guan WJ, Ni ZY, Hu Y, et al. Clinical Characteristics of Coronavirus Disease 2019 in China. N Engl J Med 2020.

5. REUTERS. Coronavirus Median Incubation Period 5-7 Days, Maximum 14: Chinese Medical Association. 2020:https://www.reuters.com/article/us-health-coronavirus-china/coronavirus-median-incubation-period-5-7-days-maximum-14-chinese-medical-association-idUSKBN20R14D.

6. Nishiura H. Determination of the appropriate quarantine period following smallpox exposure: an objective approach using the incubation period distribution. Int J Hyg Environ Health 2009;212:97–104.

7. Shira D. China’s Extended Lunar New Year Holiday Schedule. 2020:https://www.china-briefing.com/news/china-extends-lunar-new-year-holiday-february-2-shanghai-february-9-contain-coronavirus-outbreak/.

8. Chowell G, Abdirizak F, Lee S, et al. Transmission characteristics of MERS and SARS in the healthcare setting: a comparative study. BMC Med 2015;13:210.

9. Al-Tawfiq JA. Asymptomatic coronavirus infection: MERS-CoV and SARS-CoV-2 (COVID-19). Travel Med Infect Dis 2020:101608.

10. Wang D, Hu B, Hu C, et al. Clinical Characteristics of 138 Hospitalized Patients With 2019 Novel Coronavirus-Infected Pneumonia in Wuhan, China. JAMA 2020.

11. Chen N, Zhou M, Dong X, et al. Epidemiological and clinical characteristics of 99 cases of 2019 novel coronavirus pneumonia in Wuhan, China: a descriptive study. Lancet 2020;395:507–13.

